# Efficacy and harms of remdesivir for the treatment of COVID-19: a systematic review and meta-analysis

**DOI:** 10.1101/2020.05.26.20109595

**Authors:** Alejandro Piscoya, Luis Fernando Ng-Sueng, Angela Parra del Riego, Renato Cerna-Viacava, Vinay Pasupuleti, Yuani M. Roman, Priyaleela Thota, C. Michael White, Adrian V. Hernandez

**Affiliations:** Unidad de Revisiones Sistemáticas y Meta-análisis (URSIGET), Vicerrectorado de Investigación, Universidad San Ignacio de Loyola (USIL), Lima, Peru (Piscoya, Ng-Sueng, Hernandez); Hospital Guillermo Kaelin de La Fuente, Lima, Peru (Piscoya); Escuela de Medicina, Universidad Peruana de Ciencias Aplicadas (UPC), Lima, Peru (Parra del Riego, Cerna-Viacava); MedErgy Health Group Inc., Yardley, PA (Pasupuleti); Health Outcomes, Policy, and Evidence Synthesis (HOPES) Group, University of Connecticut School of Pharmacy, Storrs, CT (Roman, White, Hernandez); Hemex Health Inc., Portland, OR (Thota)

## Abstract

**Background:** We evaluated the efficacy and safety of remdesivir for the treatment of COVID-19.

**Methods:** Systematic review in five engines, pre-print webpages and RCT registries until May 22, 2020 for randomized controlled trials (RCTs) and observational studies evaluating remdesivir on confirmed, COVID-19 adults with pneumonia and/or respiratory insufficiency. Primary outcomes were all-cause mortality, clinical improvement or recovery, need for invasive ventilation, and serious adverse events (SAE). Secondary outcomes included length of hospital stay, progression of pneumonia, and adverse events (AE). Inverse variance random effects meta-analyses were performed.

**Results:** Two placebo-controlled RCTs (n=1300) and two case series (n=88) were included. All studies used remdesivir 200mg IV the first day and 100mg IV for 9 more days, and followed up until 28 days. Wang et al. RCT was stopped early due to AEs; ACTT-1 was preliminary reported at 15-day follow up. Time to clinical improvement was not decreased in Wang et al. RCT, but median time to recovery was decreased by 4 days in ACTT-1. Remdesivir did not decrease all-cause mortality (RR 0.71, 95%CI 0.39 to 1.28) and need for invasive ventilation at 14 days (RR 0.57, 95%CI 0.23 to 1.42), but had fewer SAEs (RR 0.77, 95%CI 0.63 to 0.94). AEs were similar between remdesivir and placebo arms. Risk of bias ranged from some concerns to high risk in RCTs.

**Interpretation:** There is paucity of adequately powered and fully reported RCTs evaluating effects of remdesivir in adult, hospitalized COVID-19 patients. Remdesivir should not be recommended for the treatment of severe COVID-19.

## Introduction

Coronavirus Disease 2019 (COVID-19), has caused millions of infections and hundreds of thousands of deaths worldwide.^1^ COVID-19 has more severe manifestations in older populations with chronic diseases such as obesity, hypertension, diabetes, and chronic kidney disease.^2^ In the absence of an effective vaccine, medication to reduce the severity of COVID-19 manifestations is desperately needed. While many drugs are under investigation, no drugs have demonstrated efficacy in randomized controlled trials (RCTs) with a reasonable safety profile.^3^

The nucleotide analogue, remdesivir, inhibits RNA polymerase limiting viral replication.^4,5^ It was originally developed to treat Ebola but promising in vitro effects were not translated into acceptable clinical efficacy. Remdesivir has shown antiviral effects on coronaviruses in vitro and a recent study in monkeys infected with COVID-19 showed that remdesivir significantly reduced pulmonary damage when administered early.^4,6^ On May 1^st^ 2020, the Food and Drug Administration (FDA) issued an Emergency Use Authorization (EUA) for remdesivir in severe, hospitalized COVID-19 patients; valid for one year.^7^ Giving this promising designation, we systematically evaluated the human studies assessing the efficacy and safety of remdesivir for the treatment of COVID-19.

## Methods

### Search strategy and selection criteria

We performed a systematic review of RCTs and observational studies (cohort studies, case series) evaluating the effects of remdesivir in adult hospitalized COVID-19 confirmed patients. A comprehensive literature search was conducted in PubMed, Web of Science, Scopus, Embase and the Cochrane Library. The search was broadened for preprints at medRxiv.org, and for records of ongoing remdesivir RCTs at www.ClinicalTrials.gov,www.who.int/ictrp/about/en/, and www.clinicaltrialsregister.eu/. A special twitter search was performed using the advanced search setting. Databases were searched on May 5^th^, 2020 and updated on May 22^nd^, 2020.

Search strategies were adjusted for each engine using the following combination of keywords: remdesivir AND (COVID-19 OR coronavirus OR coronavirus disease OR coronavirus disease-19 OR severe acute respiratory syndrome OR SARS-CoV-2) with no limitations for time or language. The PubMed strategy is included in the Supplement. Included studies specified remdesivir dosage and duration with at least one efficacy or harm outcome. We excluded studies with the following characteristics: patients <18 years old, patients with hepatitis B or HIV coinfection, and studies published only as meeting abstracts, pre-marketing clinical trials, or duplicates.

### Data extraction

Three reviewers (VP, AP, LNS) performed searches in engines and websites and collected records in www.myendnoteweb.com and www.dropbox.com. Two independent reviewers (APdR, RCV) assessed titles and abstracts for eligibility according to the inclusion and exclusion criteria. Discrepancies were resolved by discussion among reviewers. Three independent reviewers (LNS, APdR, RCV) extracted data with disagreements and duplicate data resolved by a third reviewer (AP). Extracted information included: study authors, year of publication, study design, number of patients, country where study was conducted, median age, proportion of males, comorbidities (obesity, hypertension, diabetes, coronary artery disease, chronic kidney disease, chronic obstructive pulmonary disease), PCR method for COVID-19 diagnosis, remdesivir dose and duration, control dose and duration, concomitant treatments for both arms, primary outcomes per arm, and secondary outcomes per arm.

### Outcomes

Primary outcomes were: all-cause mortality, clinical improvement (i.e. 2-point reduction in a 6-point ordinal severity scale), time to recovery (defined as first day, during 28-day enrollment, on which a patient satisfied categories 1, 2, or 3 of an 8-point ordinal scale), need for invasive ventilation (mechanical [MV], extracorporeal membrane oxygenation [ECMO]), components of ordinal scales, and serious adverse events (SAE). Secondary outcomes were time to death, length of hospital stay, radiological progression of pneumonia, and adverse events (AE) (i.e. diarrhea, nausea, vomiting, constipation, abdominal pain, rash, hypotension, elevated liver enzymes, hyperbilirubinemia, hypoalbuminemia, low platelets, anemia, renal impairment, and coagulation impairment).

### Assessment of risk of bias

Assessment of risk of bias was performed independently by two investigators (VP, AVH) using the Cochrane RoB 2.0 tool^8^ for RCTs, and ROBINS-I tool^9^ for cohort studies with a third reviewer (AP) resolving discrepancies when needed. Case series lack a control group and cannot evaluate efficacy or harmful effects so risk of bias cannot be conducted.

### Statistical analyses

We reported our systematic review according to 2009 PRISMA guidelines.^10^ Effects of remdesivir on outcomes from individual studies were reported as hazard ratio (HR) or absolute risk difference (ARD) and their 95 confidence intervals (CIs) for dichotomous outcomes and mean differences (MD) and their 95%CIs for continuous outcomes. Inverse variance random effect meta-analyses were performed when outcome data was available for at least two studies judged to be homogeneous about study characteristics. Between study variance tau^2^ was calculated with the Paule-Mandel method. Effects of meta-analyses were reported as RRs and their 95%CIs, and heterogeneity of effects among studies was quantified with the I^2^ statistic (an I^2^>60% means high heterogeneity of effects).

### Ethics

Because this was a systematic review of published studies, no patients were involved in setting the research question or the outcome measures; thus, no ethics approval was required.

## Results

We identified 612 records in our searches. After removing duplicates, 553 articles were screened for eligibility by reviewing titles and abstracts. Among the records screened, 544 were excluded and nine full-text articles were further assessed for eligibility (Figure 1). Review of full text articles led to the exclusion of five articles due to: duplicated data (n=3), remdesivir efficacy or safety was not the main aim of study (n=1), and no outcomes for patients receiving remdesivir (n=1). Two RCTs (n=1300) and two case series (n=88) were included in the final analysis. ^11,12,13,14^ One remdesivir RCT planned to be performed in China for patients with mild and moderate COVID-19 (NCT04252664) was suspended on April 15^th^ 2020 because no eligible patients could be enrolled to the study.^15^ Characteristics of four included studies are shown in Table 1.

**Figure 1:**
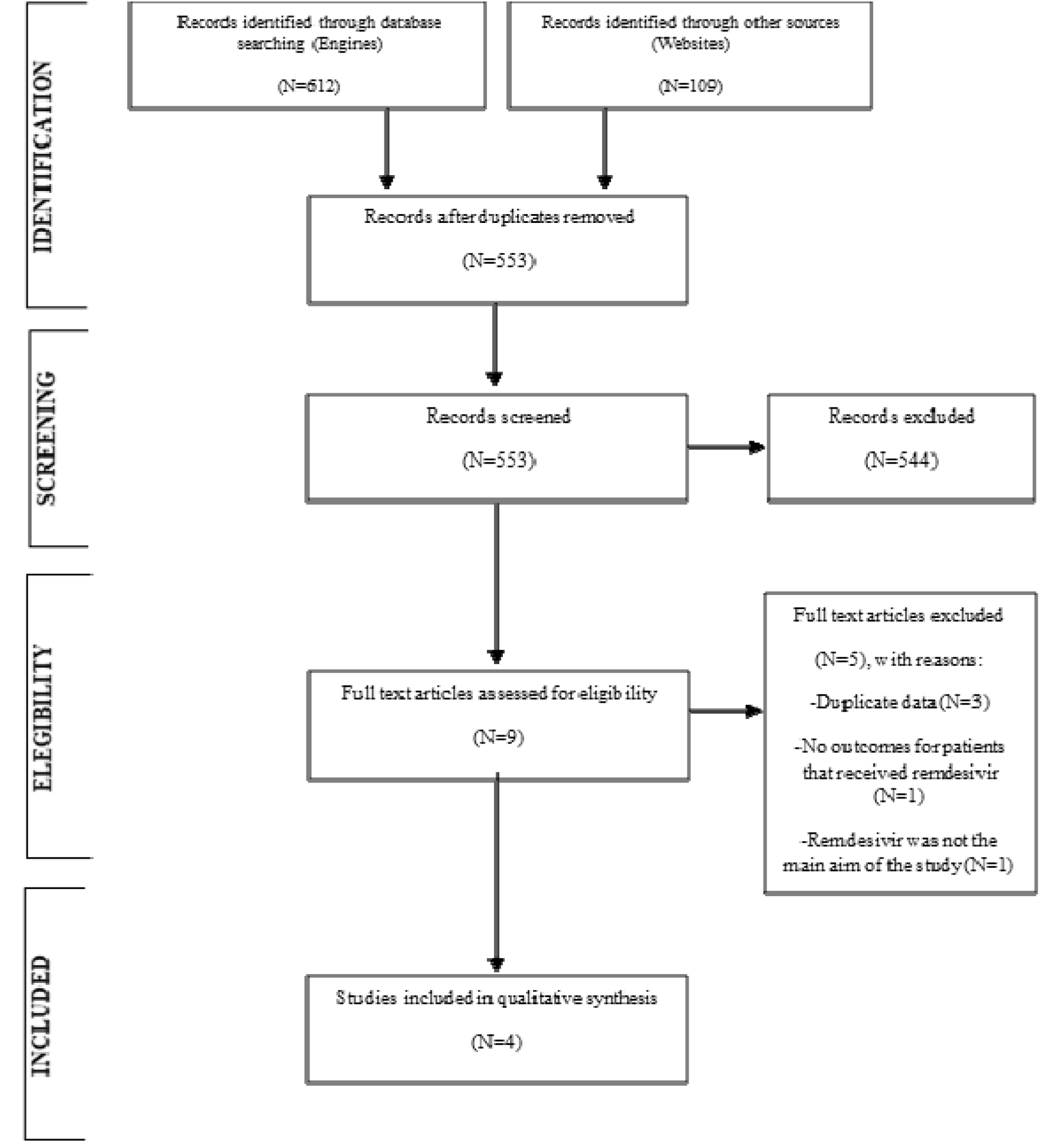
Flowchart of study selection.

**Table 1.**
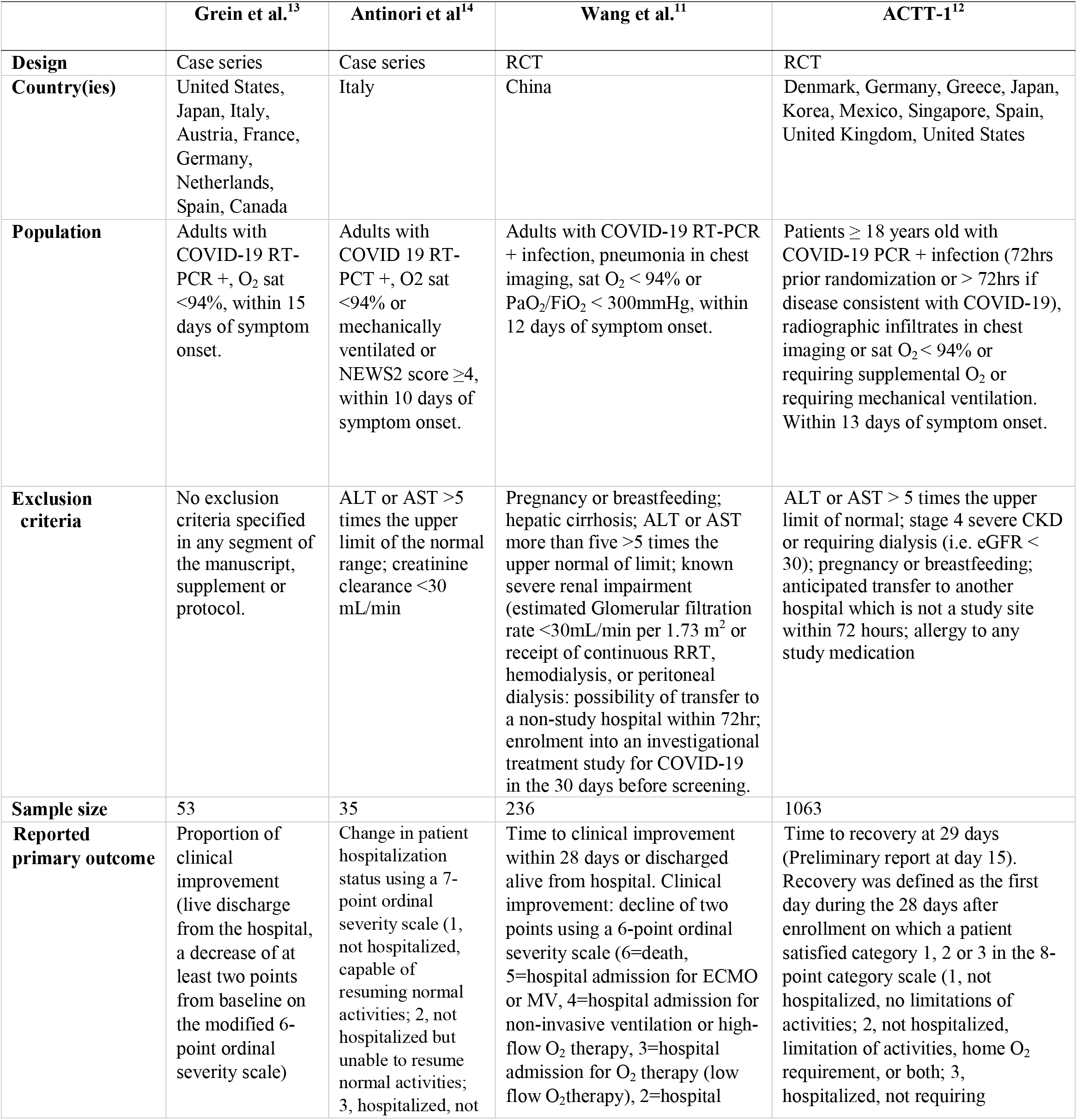
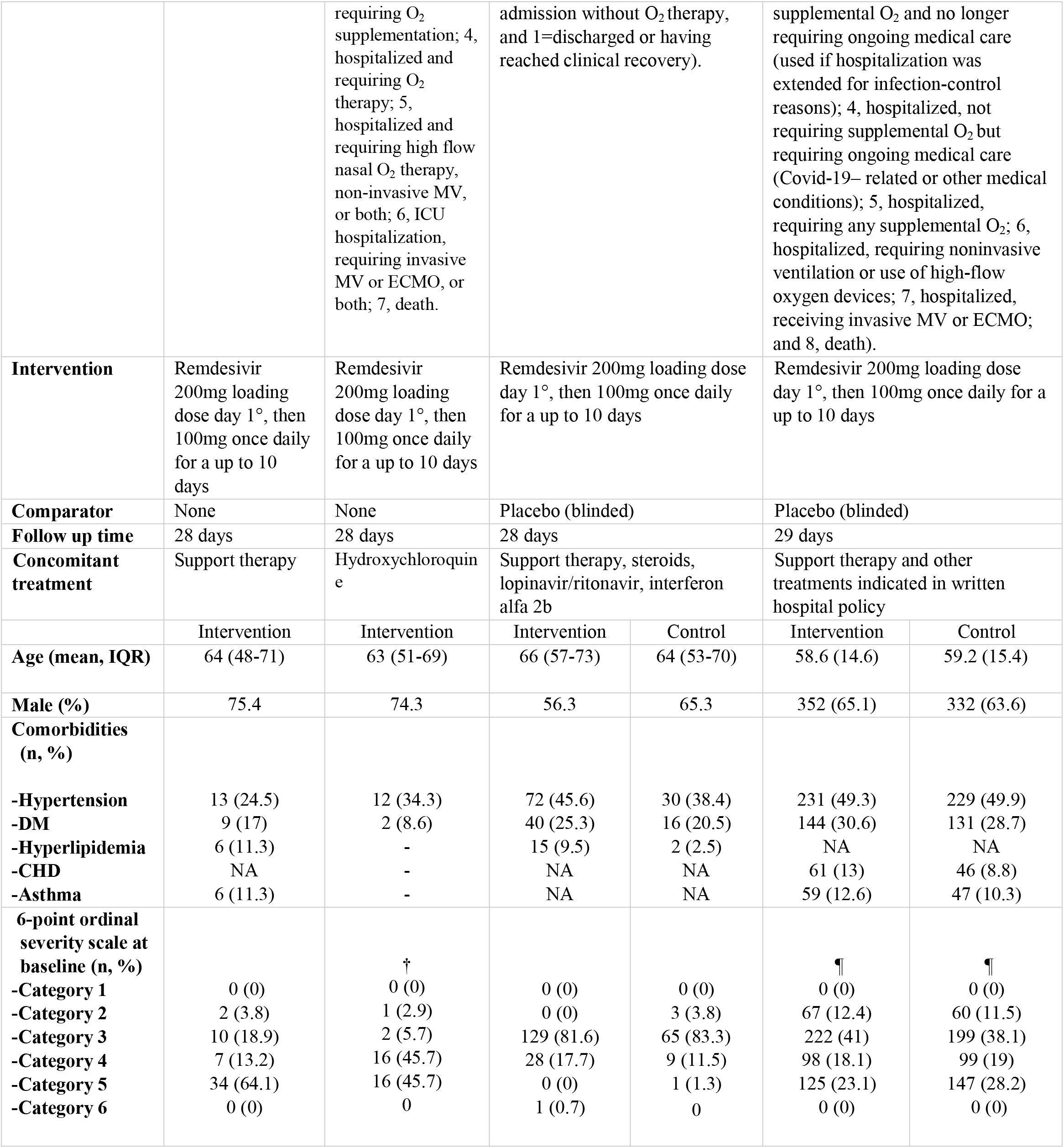
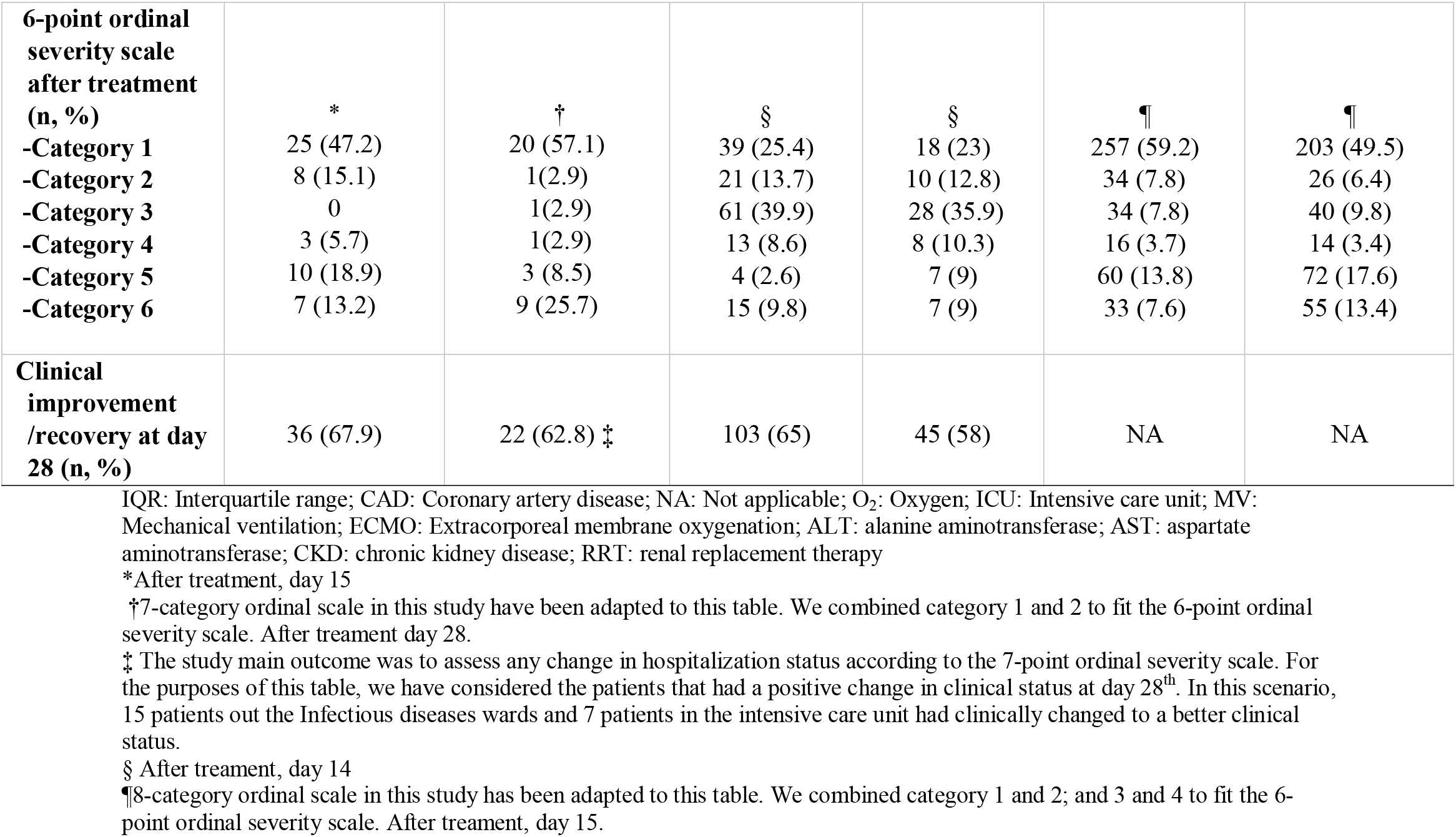
Description of characteristics of included studies

|  | Grein et al.<sup>13</sup> | Antinori et al.<sup>14</sup> | Wang et al.<sup>11</sup> | ACTT-1<sup>12</sup> |
| --- | --- | --- | --- | --- |
| **Design** | Case series | Case series | RCT | RCT |
| **Country(ies)** | United States, Japan, Italy, Austria, France, Germany, Netherlands, Spain, Canada | Italy | China | Denmark, Germany, Greece, Japan, Korea, Mexico, Singapore, Spain, United Kingdom, United States |
| **Population** | Adults with COVID-19 RT-PCR +, O<sub>2</sub> sat <94%, within 15 days of symptom onset. | Adults with COVID 19 RT-PCT +, O<sub>2</sub> sat <94% or mechanically ventilated or NEWS2 score ≥4, within 10 days of symptom onset. | Adults with COVID-19 RT-PCR + infection, pneumonia in chest imaging, sat O<sub>2</sub> < 94% or PaO<sub>2</sub>/FiO<sub>2</sub> < 300mmHg, within 12 days of symptom onset. | Patients ≥ 18 years old with COVID-19 PCR + infection (72hrs prior randomization or > 72hrs if disease consistent with COVID-19), radiographic infiltrates in chest imaging or sat O<sub>2</sub> < 94% or requiring supplemental O<sub>2</sub> or requiring mechanical ventilation. Within 13 days of symptom onset. |
| **Exclusion criteria** | No exclusion criteria specified in any segment of the manuscript, supplement or protocol. | ALT or AST >5 times the upper limit of the normal range; creatinine clearance <30 mL/min | Pregnancy or breastfeeding; hepatic cirrhosis; ALT or AST more than five >5 times the upper normal of limit; known severe renal impairment (estimated Glomerular filtration rate <30mL/min per 1.73 m<sup>2</sup> or receipt of continuous RRT, hemodialysis, or peritoneal dialysis: possibility of transfer to a non-study hospital within 72hr; enrolment into an investigational treatment study for COVID-19 in the 30 days before screening. | ALT or AST > 5 times the upper limit of normal; stage 4 severe CKD or requiring dialysis (i.e. eGFR < 30); pregnancy or breastfeeding; anticipated transfer to another hospital which is not a study site within 72 hours; allergy to any study medication |
| **Sample size** | 53 | 35 | 236 | 1063 |
| **Reported primary outcome** | Proportion of clinical improvement (live discharge from the hospital, a decrease of at least two points from baseline on the modified 6-point ordinal severity scale) | Change in patient hospitalization status using a 7-point ordinal severity scale (1, not hospitalized, capable of resuming normal activities; 2, not hospitalized but unable to resume normal activities; 3, hospitalized, not | Time to clinical improvement within 28 days or discharged alive from hospital. Clinical improvement: decline of two points using a 6-point ordinal severity scale (6=death, 5=hospital admission for ECMO or MV, 4=hospital admission for non-invasive ventilation or high-flow O<sub>2</sub> therapy, 3=hospital admission for O<sub>2</sub> therapy (low flow O<sub>2</sub>therapy), 2=hospital | Time to recovery at 29 days (Preliminary report at day 15). Recovery was defined as the first day during the 28 days after enrollment on which a patient satisfied category 1, 2 or 3 in the 8-point category scale (1, not hospitalized, no limitations of activities; 2, not hospitalized, limitation of activities, home O<sub>2</sub> requirement, or both; 3, hospitalized, not requiring |

|  |  |  |  |  |  |  |
| --- | --- | --- | --- | --- | --- | --- |
|  |  | requiring O<sub>2</sub> supplementation; 4, hospitalized and requiring O<sub>2</sub> therapy; 5, hospitalized and requiring high flow nasal O<sub>2</sub> therapy, non-invasive MV, or both; 6, ICU hospitalization, requiring invasive MV or ECMO, or both; 7, death. | admission without O<sub>2</sub> therapy, and 1=discharged or having reached clinical recovery). |  | supplemental O<sub>2</sub> and no longer requiring ongoing medical care (used if hospitalization was extended for infection-control reasons); 4, hospitalized, not requiring supplemental O<sub>2</sub> but requiring ongoing medical care (Covid-19– related or other medical conditions); 5, hospitalized, requiring any supplemental O<sub>2</sub>; 6, hospitalized, requiring noninvasive ventilation or use of high-flow oxygen devices; 7, hospitalized, receiving invasive MV or ECMO; and 8, death). |  |
| Intervention | Remdesivir 200mg loading dose day 1°, then 100mg once daily for a up to 10 days | Remdesivir 200mg loading dose day 1°, then 100mg once daily for a up to 10 days | Remdesivir 200mg loading dose day 1°, then 100mg once daily for a up to 10 days |  | Remdesivir 200mg loading dose day 1°, then 100mg once daily for a up to 10 days |  |
| Comparator | None | None | Placebo (blinded) |  | Placebo (blinded) |  |
| Follow up time | 28 days | 28 days | 28 days |  | 29 days |  |
| Concomitant treatment | Support therapy | Hydroxychloroquine | Support therapy, steroids, lopinavir/ritonavir, interferon alfa 2b |  | Support therapy and other treatments indicated in written hospital policy |  |
|  | Intervention | Intervention | Intervention | Control | Intervention | Control |
| Age (mean, IQR) | 64 (48-71) | 63 (51-69) | 66 (57-73) | 64 (53-70) | 58.6 (14.6) | 59.2 (15.4) |
| Male (%) | 75.4 | 74.3 | 56.3 | 65.3 | 352 (65.1) | 332 (63.6) |
| Comorbidities (n, %) |  |  |  |  |  |  |
| -Hypertension | 13 (24.5) | 12 (34.3) | 72 (45.6) | 30 (38.4) | 231 (49.3) | 229 (49.9) |
| -DM | 9 (17) | 2 (8.6) | 40 (25.3) | 16 (20.5) | 144 (30.6) | 131 (28.7) |
| -Hyperlipidemia | 6 (11.3) | - | 15 (9.5) | 2 (2.5) | NA | NA |
| -CHD | NA | - | NA | NA | 61 (13) | 46 (8.8) |
| -Asthma | 6 (11.3) | - | NA | NA | 59 (12.6) | 47 (10.3) |
| 6-point ordinal severity scale at baseline (n, %) |  |  |  |  |  |  |
| -Category 1 | 0 (0) | †<br>0 (0) | 0 (0) | 0 (0) | ¶<br>0 (0) | ¶<br>0 (0) |
| -Category 2 | 2 (3.8) | 1 (2.9) | 0 (0) | 3 (3.8) | 67 (12.4) | 60 (11.5) |
| -Category 3 | 10 (18.9) | 2 (5.7) | 129 (81.6) | 65 (83.3) | 222 (41) | 199 (38.1) |
| -Category 4 | 7 (13.2) | 16 (45.7) | 28 (17.7) | 9 (11.5) | 98 (18.1) | 99 (19) |
| -Category 5 | 34 (64.1) | 16 (45.7) | 0 (0) | 1 (1.3) | 125 (23.1) | 147 (28.2) |
| -Category 6 | 0 (0) | 0 | 1 (0.7) | 0 | 0 (0) | 0 (0) |
| **6-point ordinal severity scale after treatment (n, %)** |  |  |  |  |  |  |
|  | * | † | § | § | ¶ | ¶ |
| **-Category 1** | 25 (47.2) | 20 (57.1) | 39 (25.4) | 18 (23) | 257 (59.2) | 203 (49.5) |
| **-Category 2** | 8 (15.1) | 1(2.9) | 21 (13.7) | 10 (12.8) | 34 (7.8) | 26 (6.4) |
| **-Category 3** | 0 | 1(2.9) | 61 (39.9) | 28 (35.9) | 34 (7.8) | 40 (9.8) |
| **-Category 4** | 3 (5.7) | 1(2.9) | 13 (8.6) | 8 (10.3) | 16 (3.7) | 14 (3.4) |
| **-Category 5** | 10 (18.9) | 3 (8.5) | 4 (2.6) | 7 (9) | 60 (13.8) | 72 (17.6) |
| **-Category 6** | 7 (13.2) | 9 (25.7) | 15 (9.8) | 7 (9) | 33 (7.6) | 55 (13.4) |
| **Clinical improvement/recovery at day 28 (n, %)** | 36 (67.9) | 22 (62.8) ‡ | 103 (65) | 45 (58) | NA | NA |

IQR: Interquartile range; CAD: Coronary artery disease; NA: Not applicable; O<sub>2</sub>: Oxygen; ICU: Intensive care unit; MV: Mechanical ventilation; ECMO: Extracorporeal membrane oxygenation; ALT: alanine aminotransferase; AST: aspartate aminotransferase; CKD: chronic kidney disease; RRT: renal replacement therapy
\*After treatment, day 15
†7-category ordinal scale in this study have been adapted to this table. We combined category 1 and 2 to fit the 6-point ordinal severity scale. After treatment day 28.
‡ The study main outcome was to assess any change in hospitalization status according to the 7-point ordinal severity scale. For the purposes of this table, we have considered the patients that had a positive change in clinical status at day 28<sup>th</sup>. In this scenario, 15 patients out the Infectious diseases wards and 7 patients in the intensive care unit had clinically changed to a better clinical status.
§ After treatment, day 14
¶8-category ordinal scale in this study has been adapted to this table. We combined category 1 and 2; and 3 and 4 to fit the 6-point ordinal severity scale. After treatment, day 15.

### Description of studies

#### Wang et al RCT

The RCT by Wang et al (NCT04257656) assessed the efficacy and safety of intravenous (IV) remdesivir 200 mg on day one, followed by 100mg IV once-daily for nine more days vs. placebo in adults with RT-PCR confirmed SARS-CoV-2 infection, pneumonia, and respiratory insufficiency in Wuhan, China (Table 1). ^11^ The primary outcome was the time to clinical improvement within 28 days after randomization or discharged alive from the hospital, whichever came first. Clinical improvement was defined as a decline of two points using a 6-point ordinal severity scale (Table 1). This scale was modified from a 7-point ordinal severity scale used by the lopinavir/ritonavir RCT by Cao et al,^16^ which has been used in previous influenza studies by Wang. et al^17^ and recommended by the World Health Organization (WHO) R&D Blueprint expert group.^18^ This 7-point ordinal scale separated the “discharged” category into those capable of resuming normal activities or those unable to do so.

Due to higher occurrence of AEs leading to drug discontinuation vs. placebo (12% vs 5%), the trial was stopped early, with 236 recruited patients and a statistical power of 56%. Authors mentioned that they followed specific termination criteria, but the criteria are not available in the protocol.^19^ Some imbalances existed at enrollment between arms, including more patients with hypertension, diabetes, or coronary artery disease in the remdesivir arm. Also, more patients in the control group than in the remdesivir arm had been symptomatic for ≤10 days at the time of starting remdesivir or placebo treatment.

#### Beigel et al. Adaptive COVID-19 Treatment Trial (ACTT-1)

The multicenter Beigel et al. ACTT-1 RCT (NCT04280705) evaluated remdesivir 200 mg IV on day 1, followed by a 100mg IV once-daily for nine more days vs. placebo in adults with RT-PCR confirmed SARS-CoV-2 infection, pneumonia, and respiratory insufficiency (Table 1).^12, 20^ According to historical changes made to the trial at clinicaltrials.gov, on February 20^th^ 2020, its primary outcome was the percentage of subjects reporting each severity rating on the aforementioned 7-point ordinal scale from Cao et al at 15 days.^16^ On March 20^th^ 2020, the primary outcome was changed to a new 8-point ordinal severity scale, in which a subdivision was made on hospitalized patients (i.e. 5=hospitalized, not requiring supplemental oxygen - requiring ongoing medical care, and 6=hospitalized, not requiring supplemental oxygen - no longer requires ongoing medical care) (Table 1).^16^

On March 22^nd^ 2020, blinded statisticians recommended changing again the outcome to time to recovery, which was defined as the first day during the 28 days after enrollment on which a patient satisfied category 1, 2 or 3 in the 8-point category scale.^12^ This trial was stopped on April 29^th^ 2020, given that its safety monitoring board decided to release preliminary results, pointing out that the primary efficacy endpoint had been achieved.^21^ At that moment, 1063 had been recruited, and 482 recoveries (exceeding the number needed for the trial) and 81 deaths had been entered to the database.^12^ No substantial imbalances in baseline characteristics were observed between the remdesivir group and the placebo group.

#### Case series

The case series by Grein et al^13^ was conducted on a multinational basis. All 53 patients received remdesivir 200 mg IV on day one, followed by a 100mg IV once-daily for nine more days as compassionate use in RT-PCR confirmed COVID-19 patients, with oxygen saturation <94%, and a follow up of 28 days. Clinical improvement (live discharge from the hospital, a decrease of at least two points from baseline on the modified 6-point ordinal severity scale)^18^ and key clinical events (changes in oxygen support, AEs, discharge, deaths) were recorded. In this study, the mean age was 64 (48 to 71) years and 75% were male. About 60% patients had hypertension, diabetes, hyperlipidemia and asthma and most of patients were either on low flow oxygen support or invasive ventilation (i.e. categories 3 and 5 from the 6-point scale).

A second case series by Antinori et al. from Italy evaluated 35 RT-PCR confirmed COVID-19 patients, with oxygen saturation <94% or mechanically ventilated or NEWS2 score ≥4.^14^ Patients received a loading IV dose of 200 mg of remdesivir on day one and then 100 mg IV daily dose through day two to ten to assess changes on patient hospitalization status on day 10 and 28. They used the 7-point ordinal severity scale used by Cao et al.^16^ In this study, the median age was 63 (51 to 69) years with 74% male, and 9% had diabetes, 34% hypertension, 9% obesity and 3% cancer. Eighteen patients were in the ICU and 17 patients in an infectious disease ward.

#### Effect of remdesivir on primary outcomes

We merged the 6-point severity scale used in Wang et al. and the 8-point severity scale used in Beigel et al. into 5 ordinal categories (outpatient; hospitalized, non-oxygen user; hospitalized, oxygen user non-invasive; hospitalized, invasive ventilation; and death). There was no reduction of all-cause mortality in the two RCTs at 14 days (RR 0.71, 95%CI 0.39 to 1.28) (Figure 2). Grein et al reported that 13% had died^13^, whereas in Antinori et al 26% died.^14^ Time to clinical improvement was not different between arms in Wang et al. (HR 1.23, 95% CI 0.87–1.75), but time to recovery was significantly shorter with remdesivir (11 days, 95%CI 9 to 12 vs. 15 days, 95% CI 13 to 19, RR for recovery 1.32; 95%CI 1.12 to 1.55).

**Figure 2.**
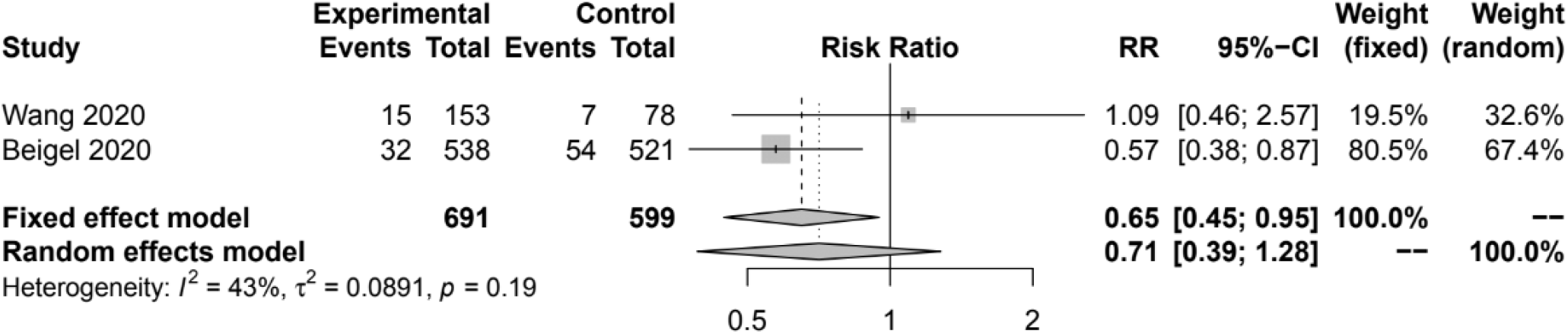
Effect of remdesivir on all-cause mortality at 14 days.

**Figure 3.**
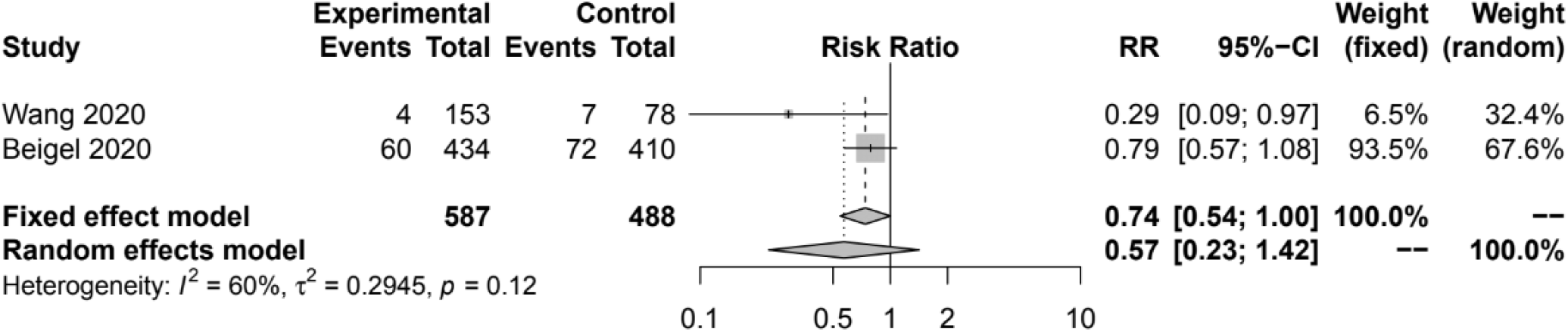
Effect of remdesivir on invasive ventilation at 14 days.

Remdesivir did not decrease the need for invasive ventilation in both RCTs (RR 0.57, 95%CI 0.23 to 1.42). SAE were significantly lower with remdesivir in the two RCTs (RR 0.77, 95%CI 0.63 to 0.94) (Figure S1). In ACCT-1, SAE were present in 21.1% vs. 27% in remdesivir and placebo, respectively. The most common SAEs were acute respiratory failure, hypotension, viral pneumonia and acute kidney injury, more frequent in the placebo arm. Grein et al. reported 23% of SAE, most commonly multiple organ failure, septic shock, kidney injury and hypotension with an 8% drug discontinuation rate.^13^

#### Effect of remdesivir on secondary outcomes

There was no effect of remdesivir in hospitalization without oxygen or with oxygen support/noninvasive ventilation in both RCTs (Figures S2 and S3). There was a higher proportion of discharged patients with remdesivir (Figure S4), and there was no different in treatment discontinuation (Figure S5). There was no difference in AEs in the two RCTs (RR 0.94, 95%CI 0.81 to 1.10) (Figure S6). No effect of remdesivir was found on specific AEs such as anemia, elevated liver enzymes, hyperbilirubinemia, hypoalbuminemia, deep vein thrombosis, pulmonary embolism or renal impairment (Figures S7 to S13).

Antinori et al. reported that 23% of AE led to drug discontinuation but with no further description.^14^ Grein et al reported 60% for AE and the most prevalent were elevated liver enzymes (23%), diarrhea (9%) and rash (8%).^13^ Antinori et al. reported elevated liver enzymes in 43% of patients, followed by acute kidney injury (23%) and elevated bilirubin levels (20%).^14^ None of the studies documented radiological progression or viral clearance at follow up.

#### Risk of bias of included RCTs

The risk of bias of each of the 2 RCTs are shown in Figure S14. Wang et al. RCT had some concerns about the comparability of arms at baseline, and ACTT-1 had high risk of bias due to selection of reporting results.

#### Ongoing trials

Table 2 describes details of six ongoing remdesivir RCTs.^22-31^ Further details of these RCTs can be found in the Supplement.

**Table 2.** List of ongoing remdesivir RCTs from trial registries

| **TRIAL ID** | **Title** | **Country** | **Type of Patient;**<br>**Remdesivir regimen** | **Comparators** | **Completion Date** |
| --- | --- | --- | --- | --- | --- |
| **IRCT20200405046953N1 (SOLIDARITY)** | Randomized trial of additional treatments for COVID-19 in hospitalized patients who are all receiving the local standard of care-Iranian SOLIDARITY multicentre trial | Iran | Hospitalized; daily infusion for 10 days (dose unspecified) | Standard of Care; Chloroquine or HCQ; Lopinavir/Ritonavir; Lopinavir/Ritonavir + IFN B-1a | Ongoing |
| **NCT04321616 (SOLIDARITY)** | The (Norwegian) NOR Solidarity Multicenter Trial on the Efficacy of Different Anti-viral Drugs in SARS-CoV-2 Infected Patients | Norway | Hospitalized; ICU; 200mg intravenous loading dose, followed by 100mg once-daily maintenance dose for 10 days | Standard of Care; HCQ | August, 2020 (Ongoing) |
| **EUCTR2020-001366-11 (SOLIDARITY)** | An international randomized trial of additional treatments for COVID-19 in hospitalized patients who are all receiving the local standard of care | Spain; Italy | Hospitalized; 100mg intravenous (duration unspecified) | HCQ; Chloroquine; Lopinavir/Ritonavir; IFN B-1a | Ongoing |
| **NCT04330690 (SOLIDARITY)** | A Multi-centre, Adaptive, Randomized, Open-label, Controlled Clinical Trial of the Safety and Efficacy of Investigational Therapeutics for the Treatment of COVID-19 in Hospitalized Patients (CATCO: Canadian Treatments for COVID-19), in Conjunction With the Public Health Emergency SOLIDARITY Trial | Canada | Hospitalized; 200mg intravenous loading dose, followed by 100mg once-daily maintenance dose for 10 days | Standard of Care; HCQ; Lopinavir/Ritonavir | March 18, 2023 (Ongoing) |
| **NCT04349410** | The Fleming [FMTVDM] Directed CoVid-19 Treatment Protocol | United States | Hospitalized; 200mg intravenous loading dose, followed by 100mg once-daily maintenance dose for 10 days | HCQ + Azithromycin; HCQ + Doxycycline; HCQ + Clindamycin; HCQ + Clindamycin + Low-Dose Primaquine; HCQ + Clindamycin + High-Dose Primaquine; Tocilizumab; Methylprednisolone; IFN A-2b; Losartan; | October 11, 2020 (Ongoing) |
|  |  |  |  | Convalescent Serum |  |
| **NCT04292730**<br>**(GILEAD)** | A Phase 3 Randomized Study to Evaluate the Safety and Antiviral Activity of Remdesivir (GS-5734™) in Participants With Moderate COVID-19 Compared to Standard of Care Treatment | United States; China; France; Germany; Hong Kong; Italy; Japan; Korea; Netherlands; Singapore; Spain; Sweden; Switzerland; Taiwan; UK | Hospitalized; 200mg intravenous loading dose, followed by 100mg once-daily maintenance dose for 5 and 10 days | Remdesivir 5-Day Course; 10-Day Course; Standard of Care | May, 2020 (Ongoing) |
| **NCT04292899**<br>**(GILEAD)** | A Phase 3 Randomized Study to Evaluate the Safety and Antiviral Activity of Remdesivir (GS-5734™) in Participants With Severe COVID-19 | United States; China; France; Germany; Hong Kong; Italy; Japan; Korea; Netherlands; Singapore; Spain; Sweden; Switzerland; Taiwan; UK | Hospitalized; 200mg intravenous loading dose, followed by 100mg once-daily maintenance dose for 5 and 10 days | Remdesivir 5-Day Course; 10-Day Course | May, 2020, (Ongoing) |
| **NCT04315948**<br>**(DisCoVeRy)** | Multi-centre, Adaptive, Randomized Trial of the Safety and Efficacy of Treatments of COVID-19 in Hospitalized Adults | France; Luxembourg | Mechanical Ventilation; 200mg intravenous loading dose, followed by 100mg once-daily maintenance dose for 10 days | Standard of Care; HCQ; Lopinavir/Ritonavir; Lopinavir/Ritonavir + IFN B-1a | March, 2023 (Ongoing) |
| **EUCTR2020-001052-18**<br>**(ACTT UK-EU)** | A Multicenter, Adaptive, Randomized Blinded Controlled Trial of the Safety and Efficacy of Investigational Therapeutics for the Treatment of COVID-19 in Hospitalized Adults - Version for European Union/United Kingdom Sites | Denmark; UK; Greece; Spain | Hospitalized; Mechanical Ventilation; 100 to 200mg intravenous (duration unspecified) | Placebo | Ongoing |

## Interpretation

In adult, hospitalized, RT-PCR confirmed COVID-19 patients with respiratory insufficiency or pneumonia, there was scarce data on efficacy and safety associated with the use of 10-day remdesivir regimens. RCTs used a common treatment regimen and a true placebo control.

Wang et al. RCT was stopped prematurely due to excess of serious adverse events causing drug discontinuation, and ACTT-1 was preliminarily reported at 15-days. While Wang et al described the use of a priori protocol that specified their stopping rule, there is no independent verification in their public protocol. ACTT-1 investigators changed this endpoint twice^20^ with no clear rationale for this alteration. No significant remdesivir impact on all-cause mortality, need of invasive ventilation, hospitalization without oxygen, hospitalization with oxygen or non-invasive ventilation, or treatment discontinuation was seen in RCTs. There was significantly lower incidence of SAE and higher proportion of discharged patients with remdesivir in both RCTs. There were substantial differences between the two case series in the magnitude of their outcomes.

There were some differences in the scales used to assess outcomes across studies as shown in Table 1. Wang et al. and Grein et al. used the same 6-point ordinal scale.^11,13^ Beigel et al used an 8-point scale, and Antinori et al used a 7-point scale.^12, 13^ These scales were strongly correlated, and that allowed us to formally group those scale categories in five in order to perform our meta-analyses.

According to the Emergency Use Authorization (EUA) by the FDA on May 1^st^ 2020^32^, it was reasonable to believe that remdesivir may be effective in treating COVID-19, and that, when used under the conditions described in the EUA, the known and potential benefits of remdesivir outweigh the known and potential risks of such products. This EUA allows to use remdesivir under the control of the US government, to treat adults and children with suspected or laboratory confirmed COVID-19 severe disease defined as SpO2 ≤94% on room air, requiring supplemental oxygen, MV, or ECMO, in an in-hospital setting intravenously, and in doses described in the authorized Fact Sheet.^7^ We need the 29-day follow up analyses of the ACTT in order to reach more definitive conclusions. Patients may ask specifically for remdesivir therapy because of the EUA and feel a failure to do so a substandard practice. Similarly, there is a risk that the benefits and harms of remdesivir will remain unknown if other remdesivir RCTs vs. placebo are stopped and substituted with trials where remdesivir becomes the standard of care and other experimental drugs are added onto remdesivir versus remdesivir alone.

Our systematic review has several strengths. We ran a recent and extensive systematic search in several engines and websites, and we did not restrict by language. We found commonalities across studies: all studies were in adult, hospitalized patients with severe COVID-19, in particular with pneumonia and respiratory insufficiency. All four studies evaluated the same dose and duration of remdesivir (200 mg IV the first day and then 100mg IV qd for nine more days), and the two RCTs were compared to placebo. We also systematically searched for worldwide ongoing RCTs and ongoing systematic reviews in PROSPERO that can be found in Table 3.

**Table 3.**
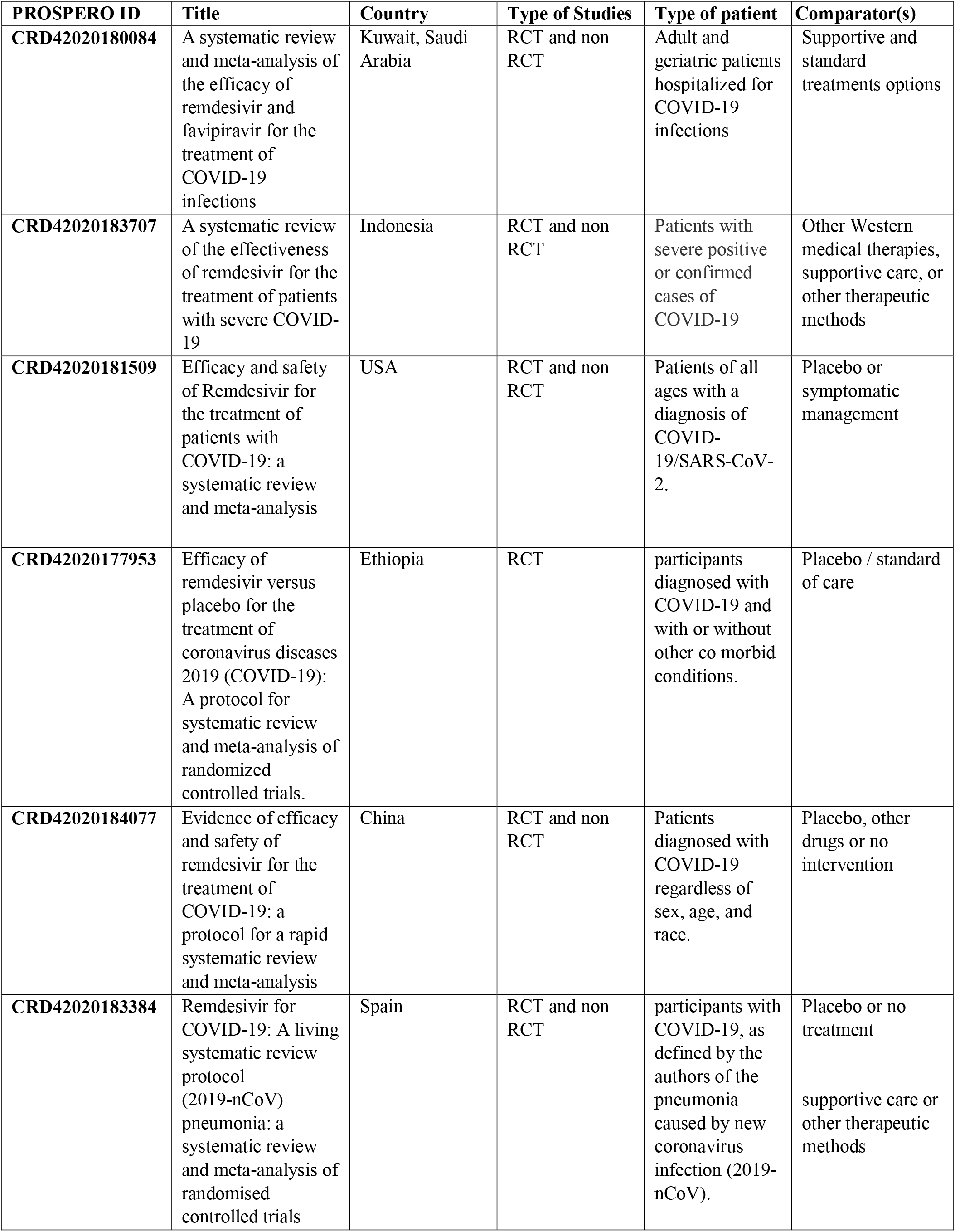
List of ongoing remdesivir systematic reviews from PROSPERO registry

| **PROSPERO ID** | **Title** | **Country** | **Type of Studies** | **Type of patient** | **Comparator(s)** |
| --- | --- | --- | --- | --- | --- |
| **CRD42020180084** | A systematic review and meta-analysis of the efficacy of remdesivir and favipiravir for the treatment of COVID-19 infections | Kuwait, Saudi Arabia | RCT and non RCT | Adult and geriatric patients hospitalized for COVID-19 infections | Supportive and standard treatments options |
| **CRD42020183707** | A systematic review of the effectiveness of remdesivir for the treatment of patients with severe COVID-19 | Indonesia | RCT and non RCT | Patients with severe positive or confirmed cases of COVID-19 | Other Western medical therapies, supportive care, or other therapeutic methods |
| **CRD42020181509** | Efficacy and safety of Remdesivir for the treatment of patients with COVID-19: a systematic review and meta-analysis | USA | RCT and non RCT | Patients of all ages with a diagnosis of COVID-19/SARS-CoV-2. | Placebo or symptomatic management |
| **CRD42020177953** | Efficacy of remdesivir versus placebo for the treatment of coronavirus diseases 2019 (COVID-19): A protocol for systematic review and meta-analysis of randomized controlled trials. | Ethiopia | RCT | participants diagnosed with COVID-19 and with or without other co morbid conditions. | Placebo / standard of care |
| **CRD42020184077** | Evidence of efficacy and safety of remdesivir for the treatment of COVID-19: a protocol for a rapid systematic review and meta-analysis | China | RCT and non RCT | Patients diagnosed with COVID-19 regardless of sex, age, and race. | Placebo, other drugs or no intervention |
| **CRD42020183384** | Remdesivir for COVID-19: A living systematic review protocol | Spain | RCT and non RCT | participants with COVID-19, as defined by the authors of the trials | Placebo or no treatment |
| **CRD42020168406** | Remdesivir for novel coronavirus | China | RCT | Patients diagnosed with | Other Western medical therapies, |
|  | (2019-nCoV)<br>pneumonia: a<br>systematic review<br>and meta-analysis of<br>randomised<br>controlled trials |  |  | pneumonia<br>caused by new<br>coronavirus<br>infection (2019-nCoV). | supportive care or<br>other therapeutic<br>methods |

Some limitations can be highlighted. First, the number of RCTs was scarce, and the reporting of the ACTT-1 is based on 15-day outcomes of the totality of recruited patients. Second, Wang et al. RCT was stopped early for presence of higher proportion of adverse events leading to drug discontinuation in an unplanned interim analysis^11^.Third, our meta-analyses for primary outcomes and secondary outcomes were based only on two RCTs; however, we used outcomes similar time points of follow up and re-categorized heterogeneous ordinal outcome scales into five categories. Finally, all studies included patients within 10 to 15 days of when symptoms began. Remdesivir antiviral activity should be the highest during the first few days of active viral multiplication, as supported by a study performed in monkeys where early administration of remdesivir prevent progression to pneumonia after SARS-CoV-2 innoculation.^6^

There is paucity of adequately powered and fully reported RCTs evaluating efficacy and harms of remdesivir use in adult, hospitalized, severely-ill COVID-19 patients. One RCT was stopped early without a clear description of the reasons and the largest ACTT-1 altered their primary endpoint twice and reported 15-day outcomes only. Several ongoing RCTs should be completed despite the EUA by the FDA until May 2021 in order to determine remdesivir clinical efficacy and harm profile. At this point, remdesivir should not be recommended for the treatment of severe COVID-19.

## Data Availability

All data will be available at request to the corresponding author

## Contributors

Study concept and design: AP, AVH. Acquisition of data: AP, LNS, APdR, RC, VP, AVH. Analysis and interpretation of data: AP, VP, YMR, AVH. Drafting of the manuscript: LNS, YMR, CMW, PT, AVH. Critical revision of the manuscript for important intellectual content: AP, LNS, APdR, RC, VP, YMR, CMW, PT, AVH. Statistical analysis: VP, YMR, AVH. Administrative, technical, and material support: AVH. Study supervision: AP, AVH. All authors approved the final manuscript as submitted. The corresponding author attests that all listed authors meet authorship criteria and that no others meeting the criteria have been omitted.

## Data sharing

All included studies are publicly available. Extracted data and risk of bias assessment files are available as web supplements.

